# Serum Antibody Fingerprinting for SARS-CoV-2 Variants in Infected and Vaccinated Subjects by Label-Free Microarray Biosensor

**DOI:** 10.1101/2023.11.02.23297831

**Authors:** Thomas Carzaniga, Luca Casiraghi, Giovanni Nava, Giuliano Zanchetta, Tommaso Inzani, Marcella Chiari, Valentina Bollati, Sara Epis, Claudio Bandi, Alessia Lai, Gianguglielmo Zehender, Tommaso Bellini, Marco Buscaglia

## Abstract

Both viral infection and vaccination affect the antibody repertoire of a person. Here we demonstrate that the analysis of serum antibodies carries information not only on the virus type that caused the infection, but also on the specific virus variant. We developed a rapid multiplex assay providing a fingerprint of serum antibodies against five different SARS-CoV-2 variants, based on a microarray of virus antigens immobilized on the surface of a label-free reflectometric biosensor. We analyzed serum from plasma of convalescent subjects and vaccinated volunteers and extracted individual antibody profiles of both total immunoglobulin Ig and IgA fraction. We found that Ig level profiles were strongly correlated with the specific variant of infection or vaccination and that vaccinated subjects displayed larger quantity of total Ig and lower fraction of IgA relative to the population of convalescent unvaccinated subjects.

## 1 INTRODUCTION

The easy access to individual antibody repertoire, which results from a complex interplay of factors Wine et al. (2015), would constitute an important achievement in providing epidemiological information, controlling disease outbreaks and developing effective clinical therapeutics and vaccine strategies Ionov and Lee (2022). During the Covid-19 pandemic, quantitative measurement of SARS-CoV-2 antibody titer enabled assessing variability in the immune response to infection, evaluating vaccine efficacy and potential for long-term immunity, as well as identifying donors for blood transfusion therapy Byrnes et al. (2020), Johnson et al. (2020), Castillo-Olivares et al. (2021), Siracusano et al. (2021), Wei et al. (2021), Ong et al. (2021), Harvey et al. (2021), Garcia-Beltran et al. (2021). Large-scale antibody quantification and characterization are commonly accomplished using Enzyme-Linked Immunosorbent Assays (ELISA) in laboratory facilities and Lateral Flow Assay (LFA) as rapid serological test at the point of care (POC). Notably, both ELISA and LFA do not allow parallel quantification of distinct antibodies and are thus not suitable for the fingerprinting of antibody repertoire Dörschug et al. (2021), Peeling et al. (2020), Criscuolo et al. (2020).

Multiplexed antigen assay platforms represent a key development for the accurate identification of antibody repertoires Ripperger et al. (2020). A high-throughput approach is offered by peptide micro-arrays, which enable identifying immunoreactive epitopes from the blood of individuals with different histories of exposures to infective agents Weber et al. (2017), Paull and Daugherty (2018), Mishra et al. (2021), Heffron et al. (2021), Cheng et al. (2021). The relevance of this approach in serodiagnostics is, however, still to be confirmed. In the context of SARS-CoV-2, a few multiplexed antigen assay platforms have been proposed, which include fluorescence protein microarray wei Jiang et al. (2020), Berre et al. (2023), as well bead-based approaches Fink et al. (2021). Despite the validity of these methodologies, the COVID-19 pandemic experience has highlighted the critical need for affordable new assay formats that offer highly sensitive, quantitative, multiplexed, and rapid immune protection profiling.

Here, we show that it is possible to discriminate antibody repertoires in serum up to the resolution of a single SARS-CoV-2 virus variant with a simple yet sensitive and quantitative assay based on label-free readout of an antigen micro-array, without additional markers to provide the signal (e.g. colorimetric, fluorescent, chemiluminescent etc.). With the same multiplex assay, it is also possible to discriminate between vaccinated and unvaccinated subjects through their total Ig profile and IgA amount. These results are based on a multi-spot biosensing technique, the Reflective Phantom Interface (RPI) Giavazzi et al. (2013), Salina et al. (2015), that enables real-time quantification of molecular binding. Overall, the proposed antibody fingerprinting method paves the way to POC characterization of antibody repertoire against specific panels of protein antigens for purposes of either individual diagnostic or population screening.

## 2 METHODS

### 2.1 Serum samples, reagents and materials

All RBD SARS-CoV-2 spike proteins (WT-RBD, *α*-RBD, *γ*-RBD, *δ*-RBD, *o*-RBD) obtained from HEK293 human embryonic kidney immortalised cell line, were purchased from Sino Biological (Beijing, P.R.China). Nucleocapsid protein was obtained from InvivoGen (Toulouse, France product code his-sars2-n). WT-LtRBD was expressed the protozoa parasite *Leishmania tarentolae* and purified Varotto-Boccazzi et al. (2021). Trimeric spike protein HexaPro was obtained from Anton Schmitz and Günter Mayer Wrapp et al. (2020), Schmitz et al. (2021). Rabbit polyclonal antibody anti-Human IgG was obtained from Abcam (Cambridge, UK product code ab7155). Goat polyclonal antibody anti-Human IgA was obtained from Invitrogen (Rockford, USA product code SA5-10252). Wedge-like glass chips (F2 optical glass,Schott) with 5° angle, with maximum thickness of 2 mm and a size of 8 mm × 12 mm, were coated with SiO_2_ to form an anti-reflection layer of 80 nm. After ozone cleaning, the chips were dip-coated with a copolymer of dimethylacrylamide (DMA), N-acryloyloxysuccinimide (NAS), and 3-(trimethoxysilyl) propyl methacrylate (MAPS)–copoly (DMA–NAS–MAPS) called MCP2 purchased from Lucidant polymers Inc. (Sunnyvale, CA, USA) Vanjur et al. (2021). All the buffers and reagents were purchased from Sigma-Aldrich (St. Louis, MO, USA) and prepared with Milli-Q pure water. Plasma samples were obtained from healthy volunteer donors and patients at the Sacco Hospital in Milan.

### 2.2 Preparation of RPI antigen microarray cartridge

Antigen proteins and control antibody were covalently immobilized on the surface of RPI sensing chips in spots with 150–200 *μ*m diameter. Droplets of spotting buffer (PBS 1X, pH 7.4 and trehalose 50 mM) containing probe proteins at concentrations of 1 mg/ml were deposited on the chip surface by an automated, non-contact dispensing system (sciFLEXARRAYER S5; Scienion AG, Berlin, Germany). After overnight incubation, the chip surface was rinsed with blocking buffer (Tris-HCl, pH 8, 10 mM, NaCl 150 mM, ethanolamine 50 mM) and distilled water and then dried. The sensor cartridges were prepared by gluing the glass chips on the inner wall of 1 cm plastic cuvettes to form a disposable cartridge. The cartridges were stored at 4 °C before use.

### 2.3 Label-free microarray measurements

The measurements were performed using the RPI apparatus described in Salina et al. (2015), comprising LED illumination with wavelength centered at 455 nm and acquisition of images of reflected light by a CCD camera (Stingray F-145C, Allied Vision). The sensor cartridges were filled with 1.3 mL of measuring buffer. Samples spikes were performed by adding 13 *μ*L of plasma with a micro pipette. The cartridges were kept at 23 °C during the measurement through a thermalized holder, and rapid mixing of the solution was provided by a magnetic stirring bar rotating at 30 Hz. Time sequences of RPI images were acquired with at 12 fps and 60 consecutive images were averaged to provide a final set of images corresponding to 5 seconds of total acquisition time per image. After a time *t*_1_ = 1 hour of acquisition, the cuvette has been emptied and filled again with measuring buffer. Then, anti-IgA antibodies were added in solution to a final concentration of 50 nM and a second set of images was acquired for a time *t*_2_ = 1 hour.

### 2.4 Data analysis

Time sequences of RPI images of the spotted surface were analyzed by a custom MATLAB program (The MathWorks, Natick, MA, USA) to obtain the brightness of each spot as a function of time *t*, and converted into the total mass surface density of molecules *σ*(*t*). The conversion of the brightness of the RPI image pixels *u*_*s*_(*t*) into surface density is performed according to:

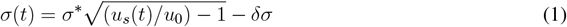

where *σ*^∗^, *u*_0_ and *δσ* are obtained as described in Salina et al. (2015) from the physical parameters of the RPI sensor, the refractive index of the solution, and the density and refractive index of a compact layer of biomolecules on the surface. The mass surface density of antibody binding the surface-immobilized antigens is obtained as Δ*σ*(*t*) = *σ*(*t*) − *σ*_0_, where *σ*_0_ is the surface density measured before the addition of plasma sample. The analysis of the binding curves was performed on Δ*σ*(*t*) traces obtained by averaging at least five spots with identical composition. Each binding curve was fitted with the exponential growth function:

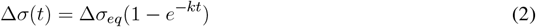

where Δ*σ*_*eq*_ is the asymptotic amplitude and *k* is the observed binding rate. The growth unit is obtained as *GU*_*Ig*_ = Δ*σ*_*eq*_*k/σ*_0_ or *GU*_*IgA*_ = Δ*σ*_*eq*_*k/*Δ*σ*(*t*_1_). *RGU* is obtained as the ratio between the *GU*_*Ig*_ of each variant RBD and that of WT-RBD.

## 3 RESULTS

### 3.1 Principle of antigen biosensor microarray

Antibody fingerprints were obtained from convalescent subjects, either vaccinated or unvaccinated, exposed to different virus variants. Plasma samples were inserted in the measuring cell (Figure 1a) hosting the RPI sensor surface (Figure 1b), which was prepared immobilizing different antigens in the form of multi-spot micro-array. The antigen panel was composed by: (i-v) five types of recombinant RBD of the spike protein of SARS-CoV-2 expressed in human cells (HEK293), corresponding to WT (WT-RBD), alpha (*α*-RBD), gamma (*γ*-RBD), delta (*δ*-RBD) and omicron (*o*-RBD) variants; (vi) full trimeric WT spike protein expressed in human cells; (vii) a variant of WT RBD expressed in Leishmania tarentolae Varotto-Boccazzi et al. (2021) (WT-LtRBD), added to evaluate the effect of different antigen glycosylation; (viii) nucleocapsid protein; (ix) antibody anti-human IgG as a positive control.

**Figure 1.**
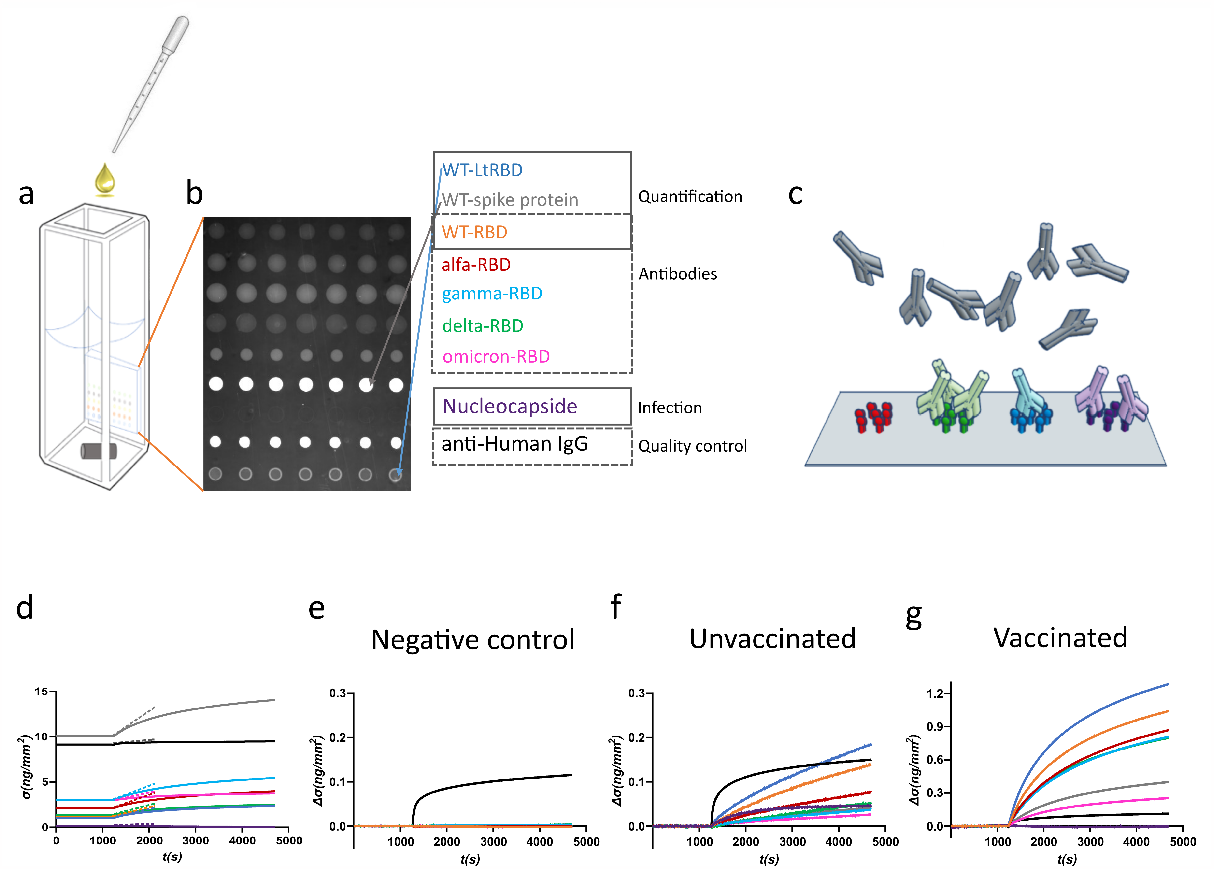
Design of the label-free microarray for anti-SARS-CoV-2 antibody fingerprinting. (a) Schematic of the plastic cartridge with the RPI sensor glued on an inner wall. Plasma samples are added to the measuring buffer with a pipette and mixing is provided by a rotating stirring bar. (b) RPI image of the sensing surface spotted with SARS-CoV-2 antigens and control proteins as indicated in the legend. (c) Cartoon of spotted sensing surface representing the binding of serum antibodies on the corresponding immobilized antigens. (d) Example of raw data of *σ*(*t*) measured before (baseline, *σ*(*t*) = *σ*_0_) and after the addition of 1:100 dilution of plasma sample. The dashed lines represent the initial slope of the binding curves. (e) Surface density Δ*σ*(*t*) = *σ*(*t*) − *σ*_0_ measured upon addition of plasma sample collected before the Covid-19 pandemic. Only the spot of anti-IgG antibody provides a signal, whereas no binding is observed on antigen spots. (f) Surface density Δ*σ*(*t*) measured upon addition of plasma sample of an unvaccinated subject previously infected by SARS-CoV-2. All antigen spots provide a positive signal but with different amplitudes. (g) Surface density Δ*σ*(*t*) measured upon addition of plasma sample of a vaccinated subject. The signal is generally larger than that obtained for convalescent subjects.

The RPI biosensor substrate consists of a wedge-shaped glass slab coated with a thin film of SiO_2_ providing anti-reflective conditions in water and with a multi-functional copolymer of dimethylacrylamide Cretich et al. (2004). When observed in the back-reflection direction, spots appear as brighter disks because the bio-conjugated proteins and the antibodies, that in time accumulate on them (Figure 1c), provide an additional effective thickness relative to the optimized anti-reflective coating condition. Images of the spotted surface (Figure 1b) were acquired before and after the addition of plasma, and the brightness of each spot was converted into surface mass density *σ* (expressed in ng/mm^2^) Salina et al. (2015). The value of *σ* reflects both the size and density of the total surface accumulation of antigens and antibodies.

Figure 1d shows an example of the assay response upon the addition of 1:100 dilution of human plasma from a vaccinated subject. *σ*_0_, the value of *σ* before the plasma addition, is larger in the spots of full spike protein and control IgG because of their larger molecular size. After the plasma addition, the molecular density on all spots increases. Figures 1e-g report three examples of different types of response observed for the increment Δ*σ*(*t*) = *σ*(*t*) − *σ*_0_ with time *t*, observed upon addition of pre-Covid-19 pandemic human plasma (Figure 1e), and of plasma from an unvaccinated convalescent subject (Figures 1f) and from a previously-uninfected vaccinated subject (Figures 1g). Pre-pandemic plasma components show negligible non-specific binding, the only significant signal is due to the presence of immunoglobulins on the anti-human IgG spot. In contrast, Δ*σ*(*t*) increases for all the antigen spots in the other two case. The differences in the responses to the different antigens between classes of subjects and within each subject enable pinpointing infection history-dependent anti-SARS-CoV-2 immunoglobulins repertoires.

### 3.2 Serum antibody fingerprint of subjects exposed to different SARS-CoV-2 variants

To quantify the relative amount of antibodies binding to the different antigens, we used the growth units *GU*_*Ig*_ determined from the growth rate of *σ*(*t*) right after the plasma addition at *t* = *t*_0_ normalized by the initial surface density of the spot: *GU*_*Ig*_ = *σ*^′^(*t*_0_)*/σ*_0_ (see Methods). The parameter *GU*_*Ig*_, being based on the slope of the linear growth of *σ*(*t*) at short times (straight lines in Fig. 1d), can be obtained with precision after a few minutes, much shorter than the time needed to estimate the asymptotic equilibrium value of the binding curve. although it provides an equivalent information. We extracted the *GU*_*Ig*_ from each antigen of each plasma sample, as well as the ratio *RGU* between the *GU*_*Ig*_ of each variant RBD and that of WT-RBD, which we adopted as an internal reference to extract accurate antibody fingerprints.

We analyzed plasma samples from 27 subjects, of which 14 were vaccinated and 13 unvaccinated and convalescent. The samples were collected between 2 and 22 days after the onset of symptoms or between 9 and 103 days after vaccination. Molecular tests identified variants of infections as WT, alfa, gamma, delta or omicron, whereas all vaccines were against WT (Supplementary Tables S1 and S2). The results obtained from a selection of samples are shown in Figure 2, where each box corresponds to plasma from a different subject and is organized in three parts. The meter on the left-hand side reports the values of *GU*_*Ig*_ relative to full spike protein (grey line), WT-RBD (orange line) and WT-LtRBD (blue line), while the one on the right-side shows the value of *GU*_*Ig*_ for the nucleocapsid. In the center, we display a radar chart (orange line) to express *RGU* for the alfa, gamma, delta and omicron RBD variants. The black line square serves as a *RGU* = 1 reference. The condition *RGU >* 1 (orange vertex outside the reference square - marked by colored circles) indicates an Ig amount larger than WT-RBD.

**Figure 2.**
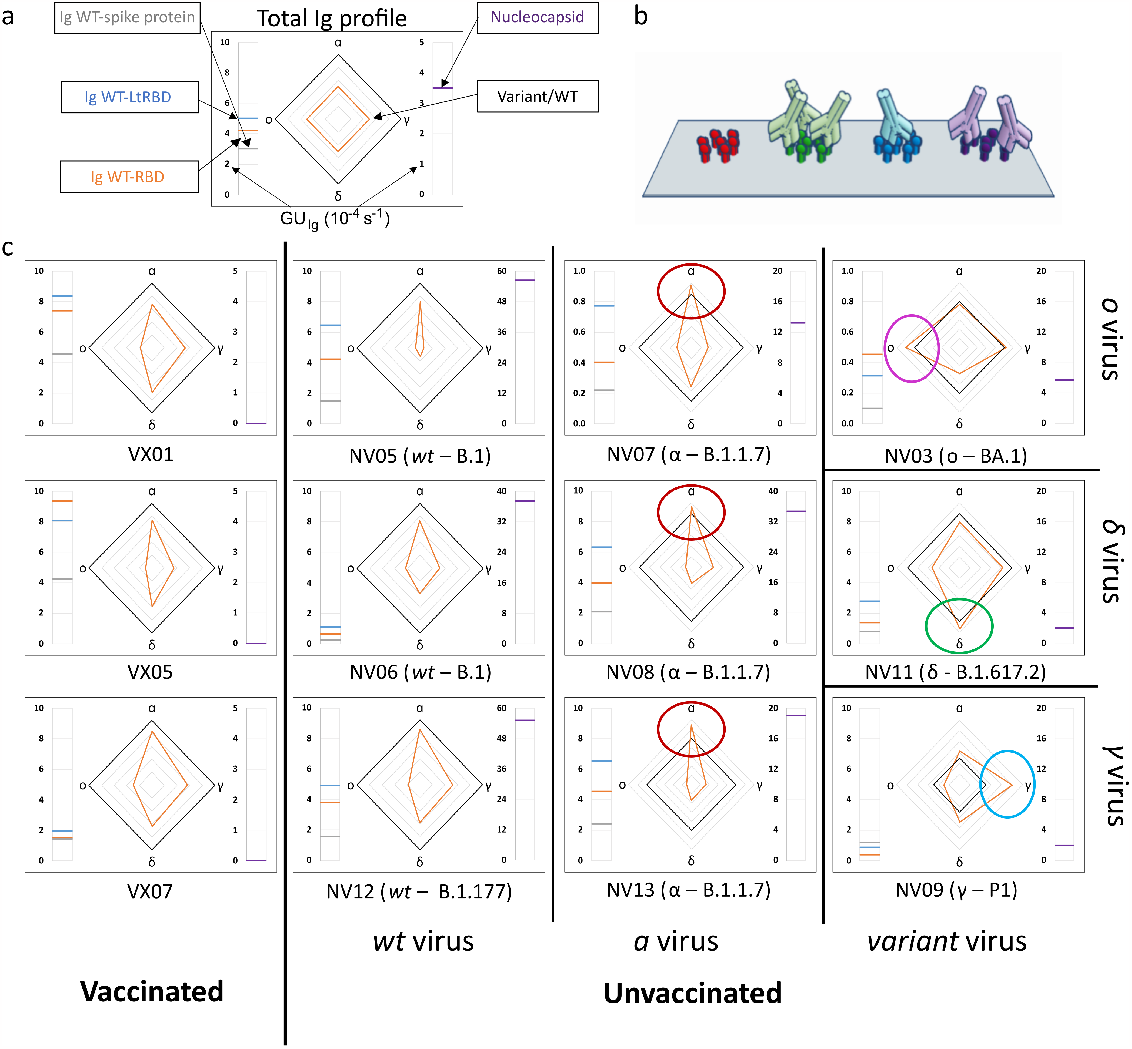
Immunoglobulins fingerprint against antigens of different SARS-CoV-2 variants. (a) Legend of the fingerprint diagram. The left-side meter reports the quantification of Ig in terms of *GU*_*Ig*_ of three WT antigens, as indicated. The right-side meter reports the quantification of anti-nucleocapsid antibodies expressed as *GU*_*Ig*_. The radar chart reports the values of *RGU* for alfa, gamma, delta and omicron RBD variants. The thick black contour line represents the amount of antibodies binding to WT-RBD taken as reference, hence corresponding to *RGU* = 1. (b) Cartoon of the assay design: Ig antibodies bind the surface-immobilized antigens. (c) Selection of Ig fingerprints obtained for three samples of vaccinated subjects (left column), and nine samples of convalescent subjects infected with different variants of of SARS-CoV-2: WT (second column from the left), alfa (third column from the left), gamma (right column, bottom), delta (right column, center), and omicron (right column, top).

Our data show a large variability among individuals of the absolute amount of Ig against the SARS-CoV-2 antigens in the sensor panel, in agreement with previous reports Wheeler et al. (2021), Siracusano et al. (2021), Wei et al. (2021). An example is provided by the meter data (left-hand side of the boxes in Figure 2), which show the absolute amount of Ig against full spike WT protein and WT RBD domains, markedly different from individual to individual. Despite this variability, a surprisingly stable pattern emerges when the ratios between Ig amounts, as expressed by RSU, are considered. This finding can be appreciated in the radar charts, where: (i) both vaccinated and unvaccinated subjects infected by the WT virus consistently display a smaller amount of Ig for the other variants (orange line always inside the black square, i.e. *RSU <* 1) with antibodies against alfa RBD always in largest amount and those targeting omicron RBD in smallest amount; (ii) Unvaccinated subjects infected by SARS-CoV-2 variants diasplay a pronounced response to the corresponding antigen, by which, for example, subjects infected by alfa variant display antibodies binding to *α*-RBD in larger amounts than to any other variants. This capacity of discriminate infection variants from the antibody response crucially relies on the multiplexing structure of our essay enabling simple computation of response ratios.

Nucleocapsid-binding Ig cannot be detected in vaccinated individuals (right-hand side meter), as expected, whereas convalescent subjects showed a variable amount of these antibodies.

The analysis of the full set of plasma samples (Supplementary Figures S1 and S2) confirms the general behaviour exemplified in Figure 2. Figure 3a-c report the pattern of relative antibodies efficiency expressed by *RGU* grouped as vaccinated, unvaccinated with past infection of WT and unvaccinated with past infection of other variants, respectively.

**Figure 3.**
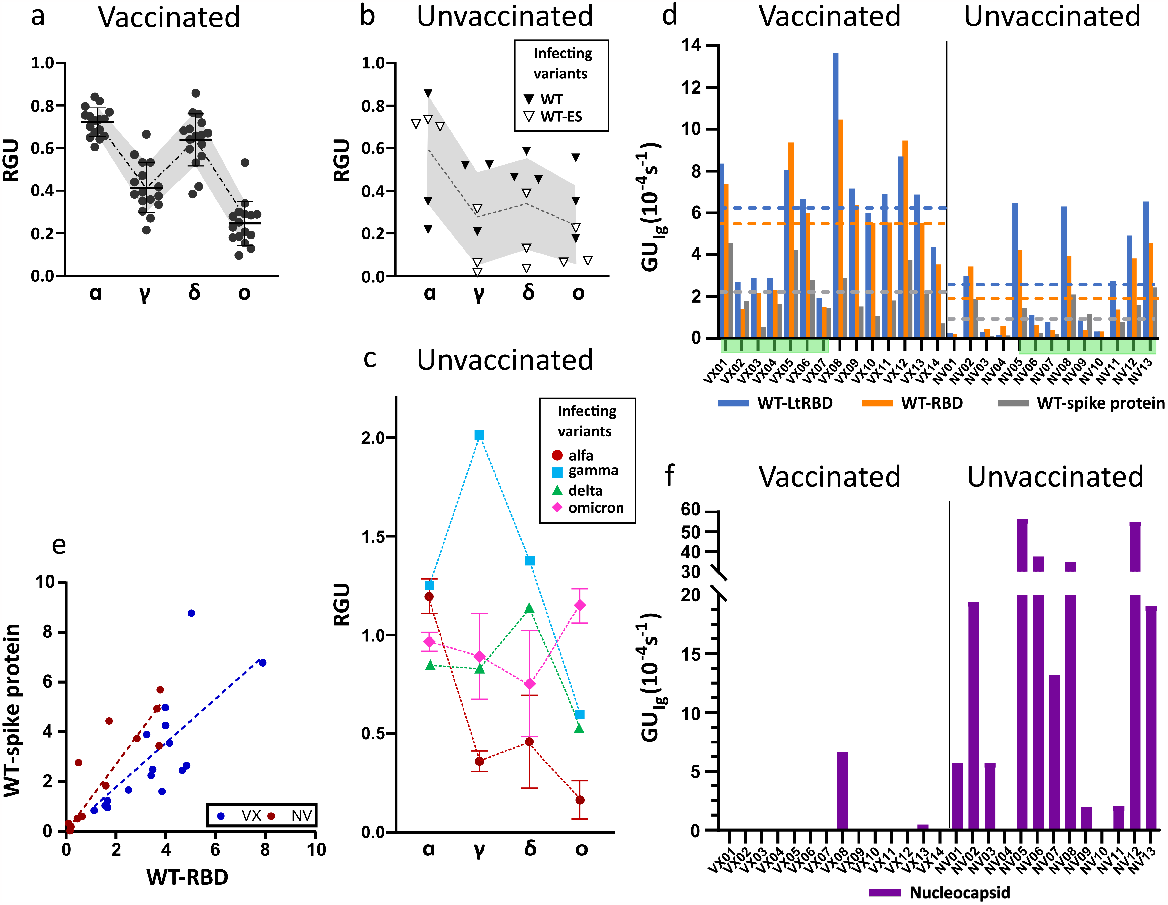
Ig fingerprint of vaccinated and unvaccinated convalescent subjects. (a) Summary of *RGU* measured for plasma samples of 14 vaccinated subjects. (b) Summary of *RGU* of 6 convalescent unvaccinated subjects infected with WT variant of strain B.1 (WT) or B.1.177 (WT-ES), as indicated in the legend. In panel a and b the dotted lines connect the average values and the borders of the shaded area connect the standard deviation values. (c) Summary of *RGU* of 7 convalescent unvaccinated subjects infected with the variants indicated in the legend. Standard deviations are indicated for variants with multiple samples. (d) Relative quantification of total Ig anti-WT spike protein and RBD, WT-RBD and WT-LtRBD, expressed as *GU*_*Ig*_ for all the analyzed samples. The dashed lines represent the average values of *GU*_*Ig*_ for all samples of vaccinated and unvaccinated subjects, the color corresponding to different antigens as indicated in the legend. (e) Growth unit *GU*_*Ig*_ of Ig on WT full spike protein and WT-RBD spots for vaccinated (blue) and unvaccinated (red) subjects. The lines represent linear fit to the data points with the corresponding color.(f) Relative quantification of anti-nucleocapsid Ig expressed as *GU*_*Ig*_ for all the analyzed samples.

All samples of vaccinated subjects (Figure 3a) display a similar pattern of response with *RGU <* 1. A similar hierarchy of binding signals is observed in samples from WT-infected convalescent subjects (Figure 3b), although in this case data are much more spread in value (grey shading). In contrast, as anticipated in Figure 2, different patterns and a larger variability of *RGU* emerge for convalescent subjects infected with the other variants (Figure 3c). In this group, for each variant of infection, the strongest response is consistently against the corresponding RBD, with values of *RGU* larger than 1.

Overall, these results demonstrate that the label-free microarray composed by 8 SARS-CoV-2 antigens can serve as antibody fingerprint accurate enough to discriminate between past infection or vaccination with different virus variants.

### 3.3 Comparison between vaccinated and unvaccinated subjects

Subjects vaccinated by WT antigen and convalescent unvaccinated subjects display *RGU* fingerprints with different features, as shown in Figure 3a-c. Further differences emerge from the analysis of the absolute quantification of Ig binding by *GU*_*Ig*_. Figure 3d shows that, on average, vaccinated subjects display larger quantities of specific antibodies against WT spike protein and RBD than unvaccinated subjects do. Data exhibit a large subject-to-subject variation coherent with the wide range of IgG concentrations estimated by ELISA, 5 - 300 ng *mL*^−1^ (Supplementary Figure S3). Figure 3d also indicates that the antibodies binding WT-LtRBD are more that those binding WT-RBD (blue vs. orange columns and lines) for both vaccinated and unvaccinated subjects, suggesting that RBD developed in human cell lines, in which glycosylation is smaller, are slightly less prone to antibody recognition. Even more significant is the difference in binding to the full spike protein (grey column and line), much weaker for vaccinated subjects in comparison to convalescent ones. This difference is also shown in Figure 3e, where it appears that, for equal response to WT-RBD, unvaccinated convalescent subjects have on average a larger response to the full spike protein.

Finally, Figure 3f shows that anti-nucleocapsid Ig are only present in samples of convalescent subjects. This is expected since SARS-CoV-2 nucleocapsid protein is not contained in vaccine formulation. Two samples of vaccinated subjects displayed anti-nucleocapsid Ig: VX08 was in prolonged contacts with infected subjects after vaccination and VX13 was presumably infected before vaccination, since some symptoms were reported. As apparent from the vertical scales in panels 3f vs. 3d, the response to nucleocapsid is extremely variable among the subjects. Indeed, while positive response to nuclecapsid is a clear indication of a previous infection, undetectable levels of anti-nucleocapsid antibodies is not necessarily an indication of the absence of previous infections, as in the case of samples NV04 and NV10, negative to nucleocapsid despite their past infection, as confirmed by molecular testing.

### 3.4 Serum immunoglobulin A fingerprint for SARS-CoV-2 exposure

The label-free assay can be enriched with the capability of discriminating between types of antibodies by measuring the binding of anti-antibodies to Ig already bound on the antigen spots after the first measuring step described above (Figure 4a). This was done by replacing the plasma in the measuring cartridge with buffer containing anti-IgA antibodies. In analogy to the quantification offered by *GU*_*Ig*_, we extracted from the data the slope of the initial linear growth of anti-IgA antibodies surface density *σ*(*t*), which we normalized to the surface density Δ*σ*(*t*_1_) of Ig at the end of the first measuring step. The resulting parameter *GU*_*IgA*_ = *σ*^′^(*t*_1_)*/*Δ*σ*(*t*_1_) (see Methods) represents the fraction of IgA in the Ig repertoire for a specific antigen.

**Figure 4.**
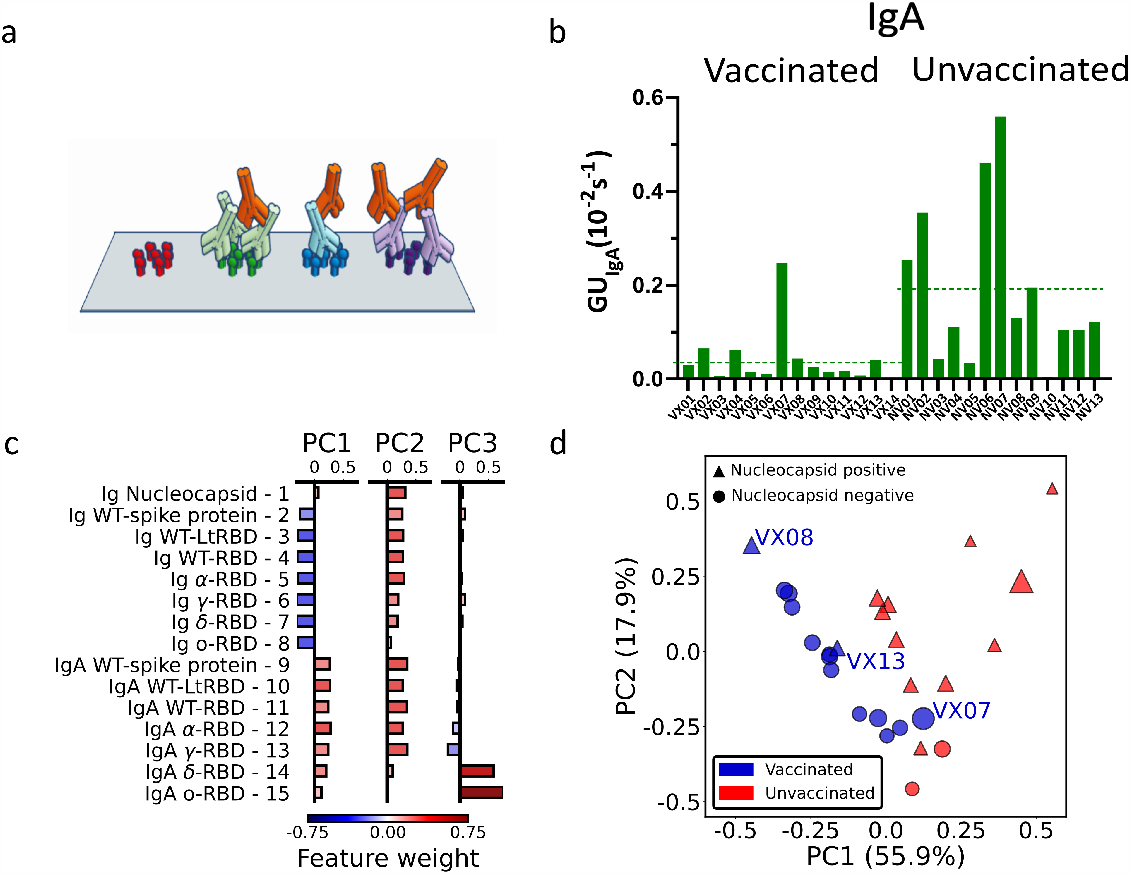
IgA fraction and principal component analysis of antibody fingerprint. (a) Cartoon of the IgA assay design: IgA antibodies (red) bind the Ig that previously bound the surface-immobilized antigen in the first step of the assay. (b) Fraction of IgA expressed as average value of *GU*_*IgA*_ on all the RBD spots for all the analyzed samples. The dashed lines represent the average values for all samples of vaccinated and unvaccinated subjects. (c) Weight of each feature in the components PC1, PC2 and PC3 obtained by principal component analysis. (d) Plot of all the 27 plasma samples in the PC1-PC2 plane. The size of each data point is proportional to the value of the PC3 component, color and shape indicate if the subject was vaccinated (blue) or unvaccinated (red), and if the nucleocapsid detection was positive (triangle) or negative (circle).

Radar charts analogous to those in Figure 2 are reported in Supplementary Figures S4 and S5. Differently from the quantification of the total Ig, we found no evident correlation of *GU*_*IgA*_ with the subject history. However, the overall fraction of IgA, estimated by the average *GU*_*IgA*_ for all RBD variants, tends to be generally larger for the samples of convalescent unvaccinated subjects as shown in Figure 4b (dashed lines). The difference in the IgA fraction is also evident when the results are grouped by virus variant (Supplementary Figure S6). Thus, IgA quantification reveals a behaviour opposite with respect to total Ig levels.

### 3.5 Principal component analysis of Ig and IgA fingerprints

The combined quantification of Ig and IgA against SARS-COV-2 spike and nucleocapsid proteins as described above measured in the 27 subjects provides a set of 15 x 27 parameters which can be combined to further enhance the discrimination the discrimination capability of the assay. To this aim, we performed a principal component analysis (PCA) of the whole data set of Ig and IgA data (Supplementary Note S1). The composition of the first three components (PC1, PC2 and PC3) are detailed in Figure 4c. As noticeable, PC1 approximately represented by the difference between IgA and Ig levels, whereas PC2 roughly represents the average amount of Ig and IgA together. Surprisingly, PC3 is only related to *GU*_*IgA*_ for delta and omicron variant.

In Figure 4d we plot PC2 vs. PC1 for all samples, PC3 being represented by the size of the symbols. As apparent, the first two principal components are effective in separating vaccinated (blue symbols) from unvaccinated (red symbols) subjects. Interestingly, the spreading of the data along the third component spontaneously provides an additional discrimination criteria for sample VX07, whose PC1 and PC2 values are instead similar to unvaccinated subjects.

We complemented PCA by performing data correlations, finding positive average correlation among Ig and among IgA and negative cross-correlation between the two groups (large total Ig are often associated to low levels if IgA), with the exception of anti-nucleocapsid Ig (Supplementary Figure S7 and S8).

The effectiveness of PCA in spontaneously discriminating groups of individuals, suggest that, in the presence of a larger set of data, our fingerprinting essay could be further strengthened by supervised analysis.

### 3.6 Evolution of immunoglobulin fingerprint of vaccinated subjects upon infection

To test the potential of the antigen array for detecting changes of antibody fingerprints of a subject over time, we analysed the *RGU* and *GU*_*IgA*_ profile of two vaccinated subjects before and after a symptomatic Covid-19 infection due to the omicron variant (Figure 5). As shown in Figure 5c-d, for both subjects, the *RGU* profiles (radar chart) are very well maintained over time despite the different amounts of total Ig (side meters). Remarkably, the infection by the omicron variant only provided a small but clearly detectable increase in the amount of Ig binding to the omicron RBD. In contrast, the IgA fraction profiles (Figure 5e-f) did not show a relative increase for the omicron variant. The effect on IgA was a large increase in the overall amount for all variants, suggesting a significant but poorly specific IgA amplification upon infection.

**Figure 5.**
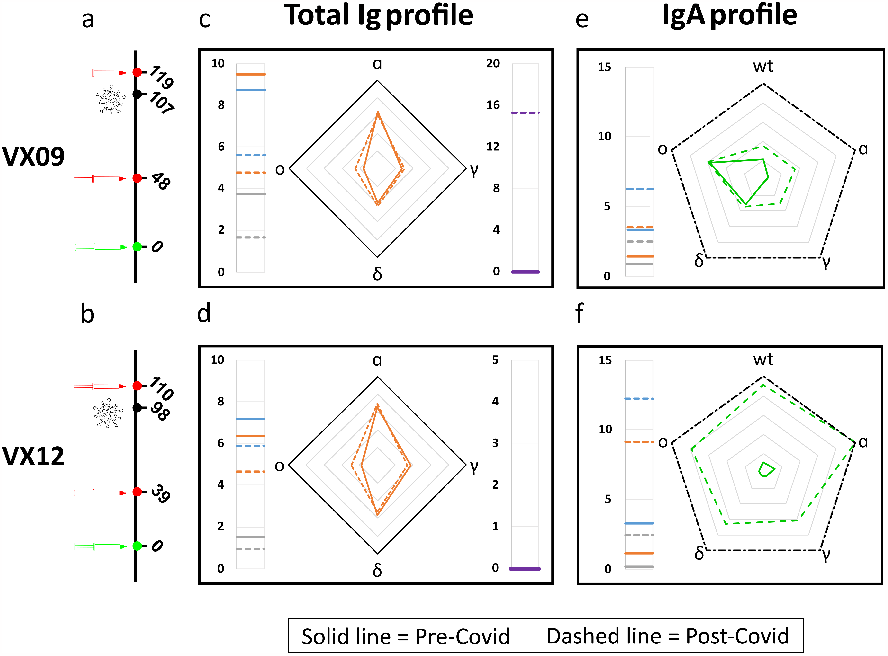
Effect of SARS-CoV-2 infection on antibody fingerprints of vaccinated subjects. (a-b) Days from vaccination (green) of sample collections (red) and infection (black) for subject VX09 (a) and VX12 (b). (c-d) Ig fingerprint (*RGU*) of subject VX09 (c) and VX12 (d) before (continuous lines) and after (dashed lines) a symptomatic infection with omicron variant. The legend for Ig fingerprints is reported in Figures 2. (e-f) IgA fraction fingerprint (*GU*_*IgA*_) of subject VX09 (e) and VX12 (f) before (continuous lines) and after (dashed lines) the infection. The left side reports the reference quantification of IgA fraction in terms of *GU*_*IgA*_ of three WT antigens: WT-spike protein (grey), WT-RBD (orange), and WT-LtRBD (blue). The radar chart reports the values of *GU*_*IgA*_ for the WT, alfa, gamma, delta and omicron RBD variants. The black dashed contour line indicates *GU*_*IgA*_ = 10^−3^*s*^−1^.

## 4 DISCUSSION

The Ig fingerprints obtained with our label-free antigen protein microarray demonstrate that the serum antibody repertoire can be analyzed up to single variant resolution, corresponding to RBD protein sequences differing by only 3 or 4 amino acids (Supplementary Figure S9). Crucial to this result is the multiple internal references offered by multiplex label-free quantification (i.e., signal background, *σ*_0_, Δ*σ*(*t*_1_), and GU of WT variant).

The RPI essay here described is primarily based on the kinetics of binding, a choice that yields an important advantage since the parameter *GU*_*Ig*_ can be quantified with a shorter measuring time than the affinity or the absolute concentration. This feature is relevant in the context of development of rapid tests suitable for POC diagnostics.

The estimated the limit of detection (LOD) of the proposed label-free micro-array, obtained by comparing the quantification of total immunoglobulins against full spike protein and WT-RBD with that of anti-WT RBD IgG measured by chemiluminescence enzyme immunoassay (Supplementary Figure S10), corresponds to 0.24 AU mL^-1^ for the antibodies against the full spike protein and 0.46 AU mL^-1^ for those against the RBD fragment, hence much lower than the typical value of 15 AU mL^-1^ considered as positive response for the chemiluminescence assay Nguyen et al. (2020).

Other novel technologies have been proposed to achieve the challenging task of combining rapidity and small LOD Cardoso et al. (2017), Calvo-Lozano et al. (2021), Zhao et al. (2021). The RPI biosensor used in this work brings the advantages of large multiplexing and cost-effective cartridge and set-up, suitable for large scale production and POC testing.

Characteristic response patterns for both Ig and IgA emerge for vaccinated and unvaccinated convalescent subjects, as confirmed by PCA analysis. On average, vaccination induces higher levels of total Ig specific to the antigen, but virus infection produces higher levels of IgA, although less antigen-specific. This behavior is confirmed also considering the time-dependence of antibody levels after virus exposure (Supplementary Figures S11 and S12). Another difference between the antibody repertoires of vaccinated and unvaccinated convalescent subjects is shown in Figure 3d, which suggests that SARS-CoV-2 infection yields a larger fraction of antibodies targeting full spike protein in regions different than RBD relative to vaccination. This is in agreement with previous work reporting a lower fraction of neutralizing antibodies in infected vs. vaccinated individuals Manenti et al. (2022).

Previous studies showed that early humoral response can be dominated by IgA antibodies, which can provide an important contribution to virus neutralization Padoan et al. (2020), Sterlin et al. (2021), Chen et al. (2020), Zervou et al. (2021), Havervall et al. (2022). Regarding the effect of vaccines on the levels of IgA, convalescent subjects were found to have larger levels of IgA than vaccinated subjects at similar time after infection or vaccination Wisnewski et al. (2021), Sano et al. (2022), Cheng et al. (2022), Sheikh-Mohamed et al. (2022). Our results are consistent with these observations and further support the difference in IgA levels between convalescent and vaccinated subjects, so that the quantification of IgA levels could be exploited for viral infection screening and virus surveillance.

In conclusion, the proposed label-free antigen microarray demonstrates the feasibility of rapid serum antibody fingerprinting discriminating among single SARS-CoV-2 variants. The results obtained suggest that this method may be useful for the serological recognition of infecting viral variants even in subjects with low viral loads or who have already eliminated the virus. This approach enables studying the epidemiology of SARS-CoV-2 infection and can be instrumental in planning strategies for control measures in the future. Our results may represent the basis for further investigations on the application of this method in contexts where it may be important to retrospectively reconstruct the infecting viral genotype in already recovered individuals or in patients with insufficient viral nucleic-acid amounts for genotyping, such as in the case of HCV or HIV or flaviviruses Murphy et al. (1999), Pawlotsky et al. (1997), Cleton et al. (2015).

## Data Availability

All data produced in the present study are available upon reasonable request to the authors

## ETHIC DECLARATION

The authors declare no competing interests. Plasma samples were collected upon approval of the Local Ethics Committee and signature of the informed consent. The ASST Fatebenefratelli Sacco, Milano, Italy, approved the protocol No. 379/2020 in March 2020, then amended in January 2023.

## AUTHOR CONTRIBUTIONS

C.B., G.Z., M.B., T.B., and V.B. designed the study; T.C. developed the analysis method and visualized data; M.C., L.C., and T.C. functionalised the chip surface; L.C. and T.C. designed and performed the experiments; G.N. and L.C. optimized the experimental setup; A.L. and G.Z provided plasma samples; C.B. and S.E. provided WT-LtRBD protein; G.N. and T.I. performed the PCA analysis; M.B., T.B, and T.C. wrote the manuscript and prepared figures with input from all of the authors.

## ACKNOWLEDGMENTS

The authors thank Anton Schmitz and Günter Mayer for the generous gift of the HexaPro spike protein. This work has received funding from Ministero dell’Universita` e della Ricerca through grants FISR 2020 COVID, “VIAEREA” project No. FISR2020IP-04633, PRIN 2017, project No. 2017Z55KCW, “Erogazione liberale per le attivita` di ricerca sul Coronavirus”, No. LIBVT20COVID19SEPIS and Fondazione “Romeo ed Enrica Invernizzi” No. LIBVT21BAND. Luca Casiraghi and Tommaso Inzani were supported by young scientist fellowships from the UNIMI GSA-IDEA project.

## SUPPLEMENTAL DATA

Supplementary tables S1-S2, supplementary figures S1-S13, and supplementary note S1.

