## Supplementary Material for "Serum Antibody Fingerprinting for SARS-CoV-2 Variants in Infected and Vaccinated Subjects by Label-Free Microarray Biosensor"

#### 1 SUPPLEMENTARY TABLES AND FIGURES

##### 1.1 Supplementary Table S1

**Table S1.** Virus strain and sample collection time for samples of convalescent subjects.

| Code | SARS-CoV-2 strain | Variant | Onset of symptoms | Plasma collection date | Days from symptoms |
|---|---|---|---|---|---|
| NV01 | B.1.177 | WT-ES | 11/2020 | 11/2020 | 2 |
| NV02 | B.1 | WT | 02/2020 | 02/2020 | 3 |
| NV03 | BA.1 | omicron 1 | 01/2022 | 01/2022 | 4 |
| NV04 | B.1.177 | WT-ES | 02/2021 | 02/2021 | 4 |
| NV05 | B.1 | WT | 02/2020 | 02/2020 | 8 |
| NV06 | B.1 | WT | 02/2020 | 02/2020 | 11 |
| NV07 | B.1.1.7 | alfa | 05/2021 | 05/2021 | 11 |
| NV08 | B.1.1.7 | alfa | 05/2021 | 05/2021 | 12 |
| NV09 | P.1 | gamma | 06/2021 | 06/2021 | 12 |
| NV10 | BA.2 | omicron 2 | 03/2022 | 03/2022 | 15 |
| NV11 | B.1.617.2 | delta | 09/2021 | 10/2021 | 17 |
| NV12 | B.1.177 | WT-ES | 03/2021 | 04/2021 | 20 |
| NV13 | B.1.1.7 | alfa | 05/2021 | 06/2021 | 22 |
| VX09b | BA.2 | omicron 2 | 04/2022 | 04/2022 | 12 |
| VX12b | BA.2 | omicron 2 | 04/2022 | 04/2022 | 12 |

.

#### 1.2 Supplementary Table S2

**Table S2.** Virus strain and sample collection time for samples of convalescent subjects.

| Code | Vaccine 1st dose | Vaccine 2nd dose | Date 2nd dose | Plasma Collection date | Days from 2nd dose |
|---|---|---|---|---|---|
| VX01 | BNT162b2 (Comirnaty) | BNT162b2 (Comirnaty) | 02/2021 | 02/2021 | 9 |
| VX02 | BNT162b2 (Comirnaty) | BNT162b2 (Comirnaty) | 02/2021 | 02/2021 | 11 |
| VX03 | Vaxzevria (ChAdOx1-S) | Vaxzevria (ChAdOx1-S) | 01/2022 | 02/2021 | 15 |
| VX04 | BNT162b2 (Comirnaty) | BNT162b2 (Comirnaty) | 02/2021 | 02/2021 | 16 |
| VX05 | BNT162b2 (Comirnaty) | BNT162b2 (Comirnaty) | 02/2021 | 02/2021 | 16 |
| VX06 | BNT162b2 (Comirnaty) | BNT162b2 (Comirnaty) | 01/2021 | 02/2021 | 20 |
| VX07 | BNT162b2 (Comirnaty) | BNT162b2 (Comirnaty) | 01/2021 | 02/2021 | 22 |
| VX08 | BNT162b2 (Comirnaty) | BNT162b2 (Comirnaty) | 01/2022 | 02/2022 | 29 |
| VX09 | Vaxzevria (ChAdOx1-S) | BNT162b2 (Comirnaty) | 12/2021 | 02/2022 | 39 |
| VX10 | Vaxzevria (ChAdOx1-S) | Vaxzevria (ChAdOx1-S) | 12/2021 | 02/2022 | 41 |
| VX11 | Vaxzevria (ChAdOx1-S) | BNT162b2 (Comirnaty) | 12/2021 | 02/2022 | 44 |
| VX12 | Vaxzevria (ChAdOx1-S) | Vaxzevria (ChAdOx1-S) | 12/2021 | 02/2022 | 48 |
| VX13 | Vaxzevria (ChAdOx1-S) | Vaxzevria (ChAdOx1-S) | 11/2021 | 02/2022 | 63 |
| VX14 | BNT162b2 (Comirnaty) | BNT162b2 (Comirnaty) | 10/2021 | 02/2022 | 103 |

.

#### 1.3 Supplementary Figure S1

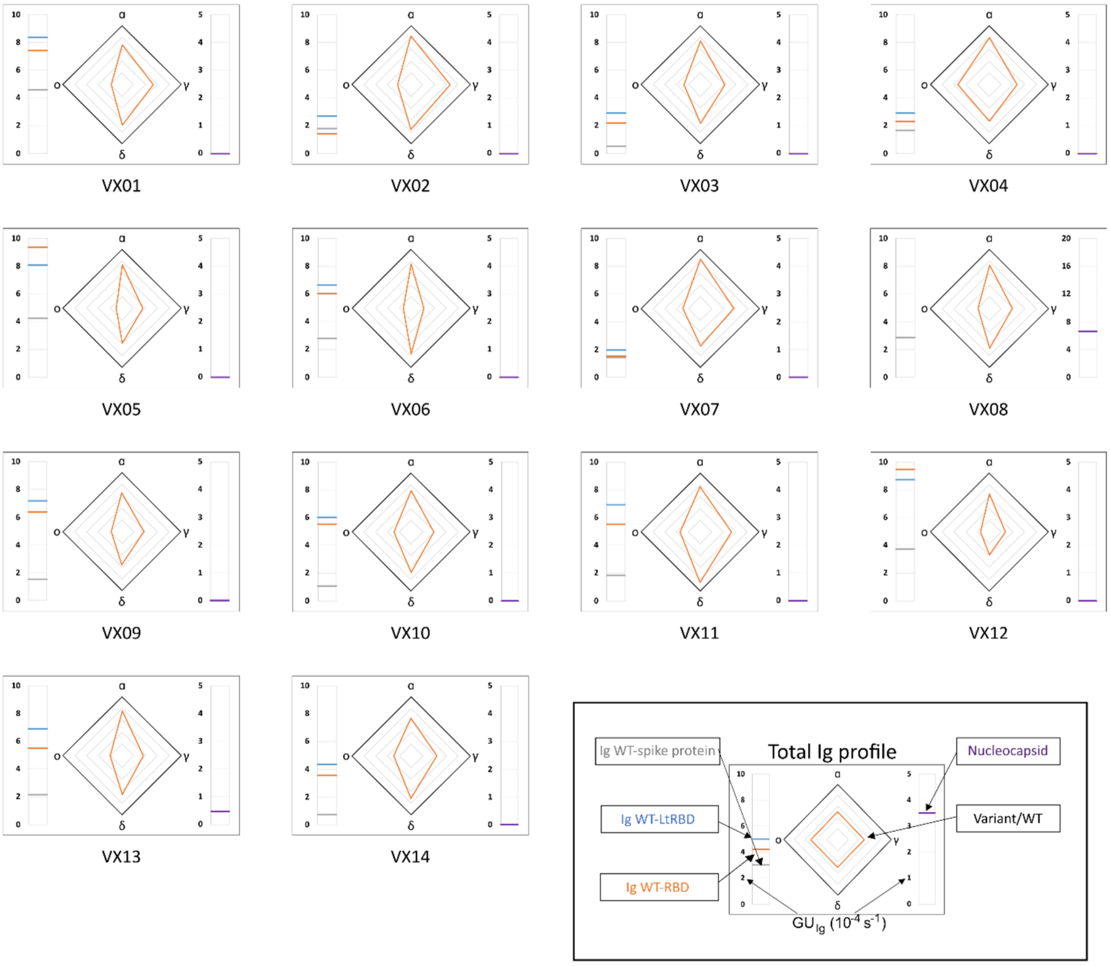


**Figure S1.** Immunoglobulins fingerprints of all samples of vaccinated subjects. The legend of the fingerprint diagram is shown in the box at the bottom of the figure. The left side reports the quantification of Ig in terms of $GU_{Ig}$ of three WT antigens, as indicated. The right side reports the quantification of anti-nucleocapsid antibodies expressed as $GU_{Ig}$. The radar chart reports the values of RGU for the alfa, gamma, delta and omicron RBD variants. The thick black contour line represents the reference amount of antibodies binding to WT-RBD hence corresponding to RGU = 1.

#### 1.4 Supplementary Figure S2

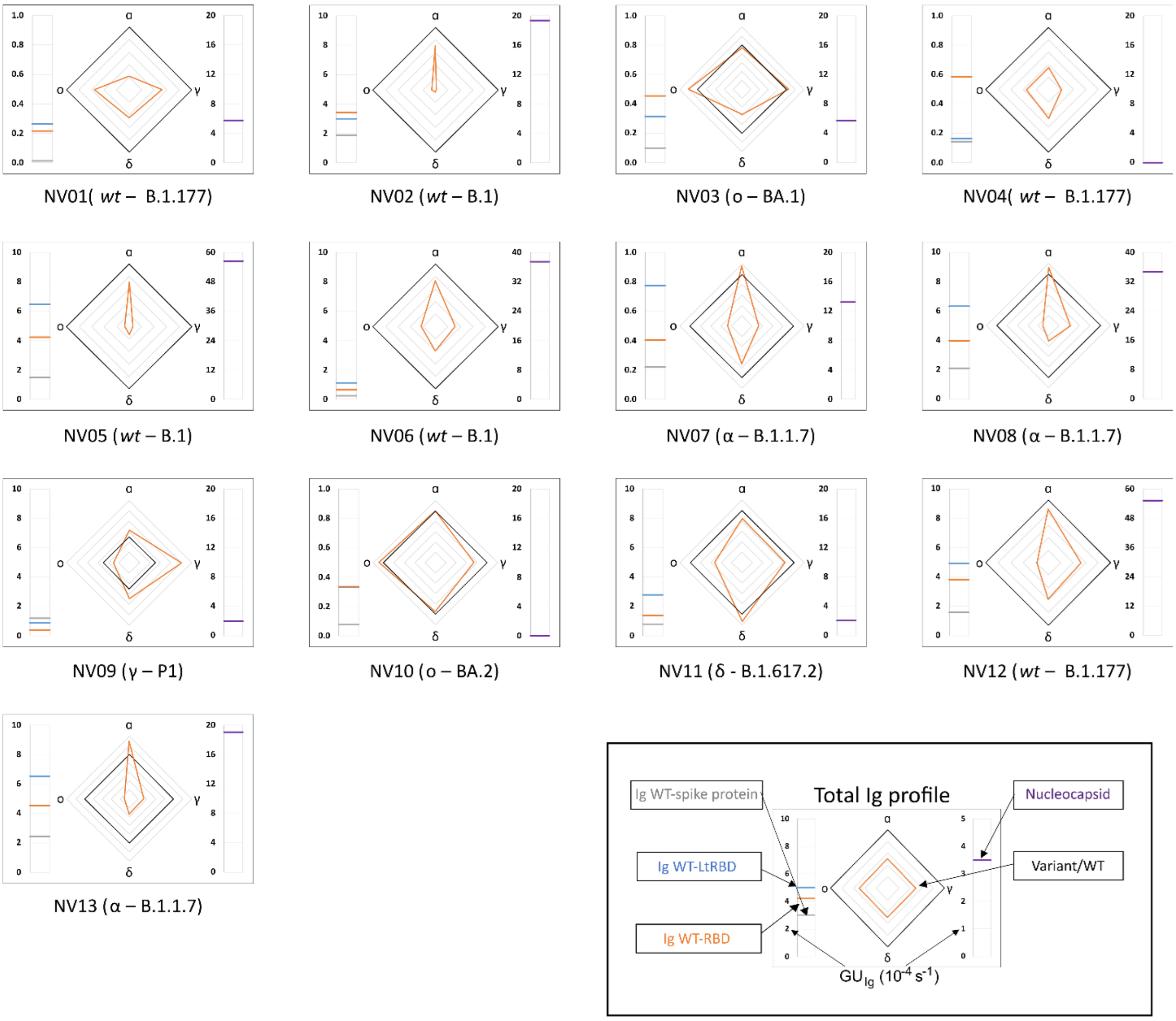


**Figure S2.** Immunoglobulins fingerprints of all samples of convalescent unvaccinated subjects. The legend of the fingerprint diagram is shown in the box at the bottom of the figure. The left side reports the quantification of Ig in terms of $GU_{Ig}$ of three WT antigens, as indicated. The right side reports the quantification of anti-nucleocapsid antibodies expressed as $GU_{Ig}$. The radar chart reports the values of RGU for the alfa, gamma, delta and omicron RBD variants. The thick black contour line represents the reference amount of antibodies binding to WT-RBD hence corresponding to RGU = 1.

#### 1.5 Supplementary Figure S3

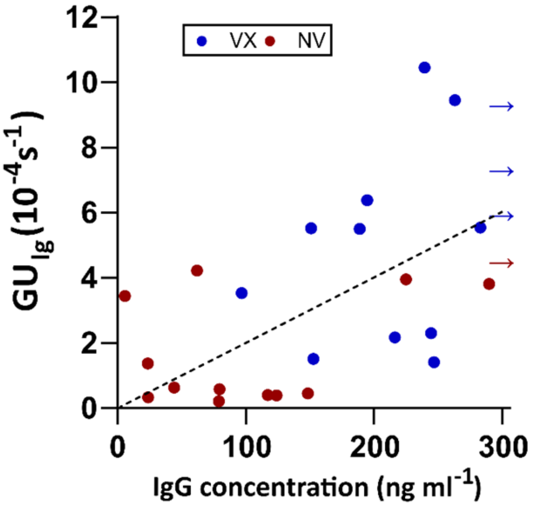


**Figure S3.** Quantification of IgG by ELISA. The growth units of total immunoglobulins obtained by label-free microarray are plotted as a function of the concentration of IgG estimated by ELISA for vaccinated (blue) and unvaccinated (red) subjects. The arrows indicate data points of $GU_{Ig}$ exceeding the saturation level for ELISA quantification. The dashed line corresponds to a linear fit with slope 0.02 $s^{-1}$/(ng $mL^{-1}$).

#### 1.6 Supplementary Figure S4

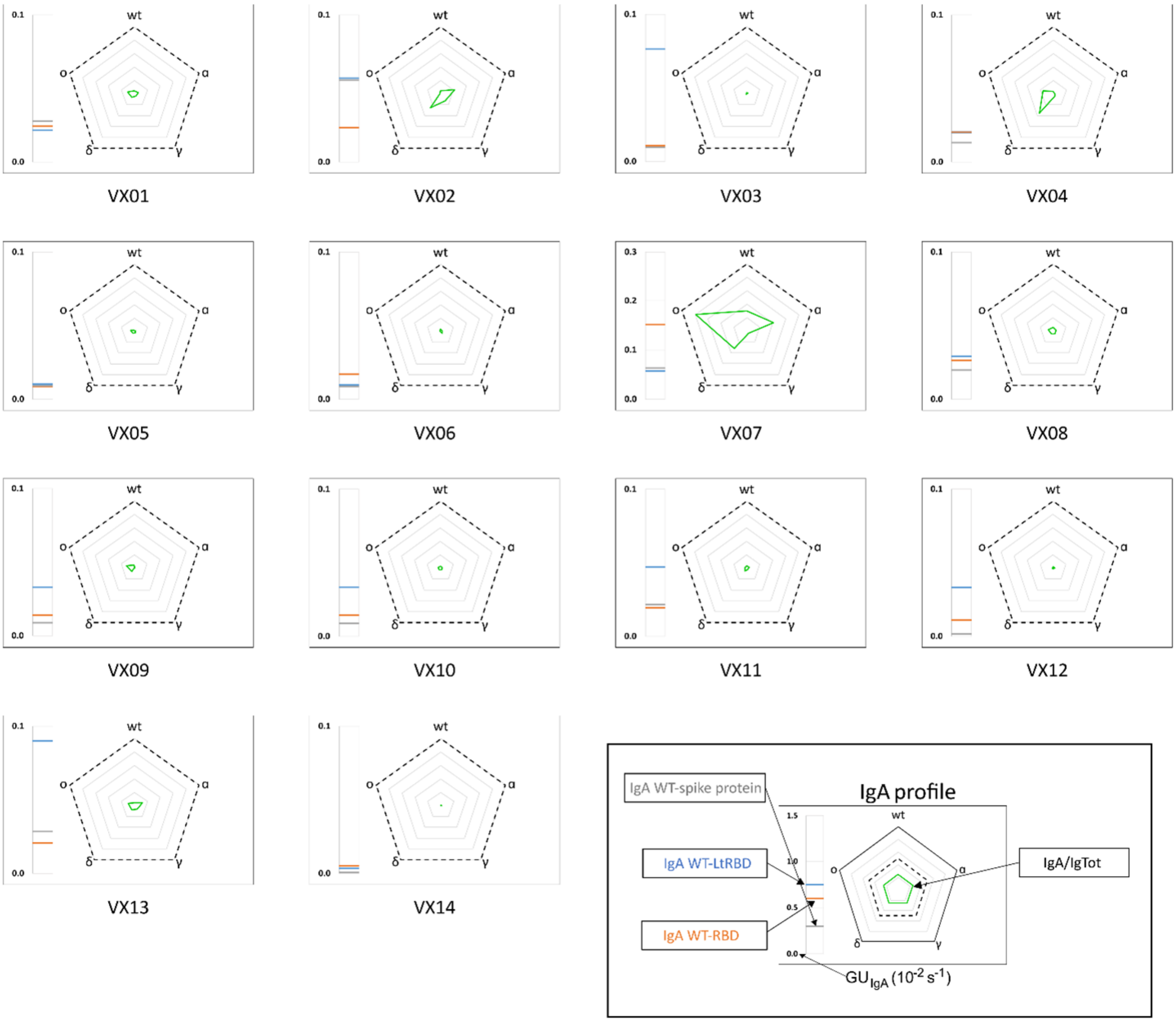


**Figure S4.** Fingerprint of IgA fraction against different SARS-CoV-2 antigen variants of all samples of vaccinated subjects. The legend of the fingerprint diagram is shown in the box at the bottom of the figure. The left side reports the reference quantification of IgA fraction in terms of $GU_{IgA}$ of three WT antigens, as indicated. The radar chart reports the values of $GU_{IgA}$ for the WT, alfa, gamma, delta and omicron RBD variants. The thick black contour line and the dashed contour line indicate $GU_{IgA} = 10^{-2}\ s^{-1}$ and $GU_{IgA} = 0.5 \cdot 10^{-2}\ s^{-1}$ , respectively.

#### 1.7 Supplementary Figure S5

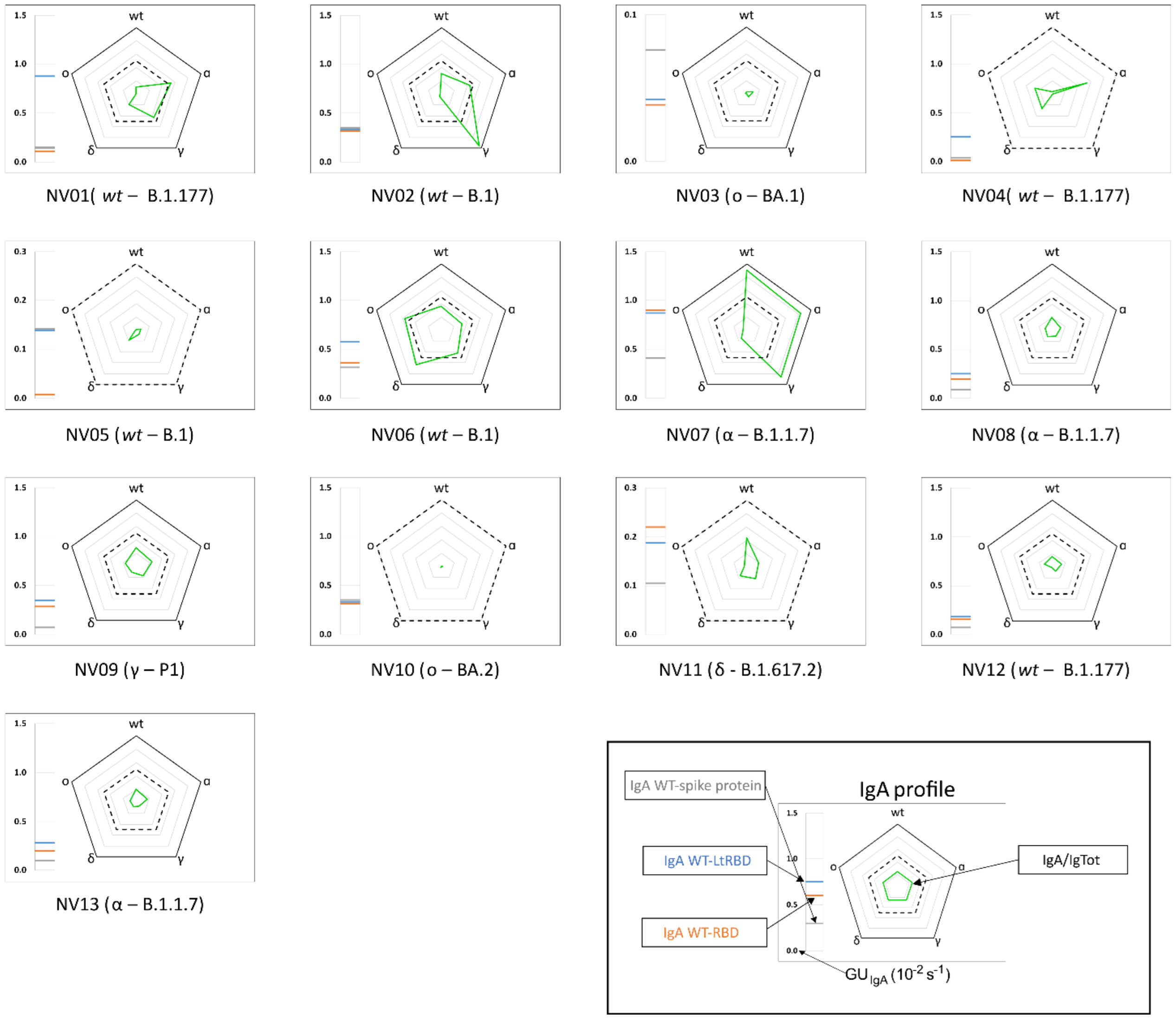


**Figure S5.** Fingerprint of IgA fraction against different SARS-CoV-2 antigen variants of all samples of convalescent unvaccinated subjects. The legend of the fingerprint diagram is shown in the box at the bottom of the figure. The left side reports the reference quantification of IgA fraction in terms of $GU_{IgA}$ of three WT antigens, as indicated. The radar chart reports the values of $GU_{IgA}$ for the WT, alfa, gamma, delta and omicron RBD variants. The thick black contour line and the dashed contour line indicate $GU_{IgA} = 10^{-2}$ $s^{-1}$ and $GU_{IgA} = 0.5 \cdot 10^{-2}$ $s^{-1}$ , respectively.

#### 1.8 Supplementary Figure S6

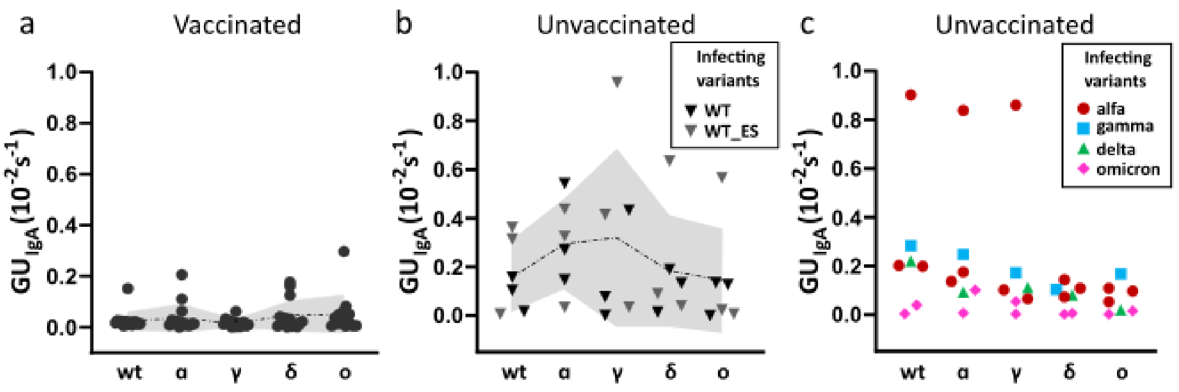


**Figure S6.** Fingerprint of IgA fraction against different SARS-CoV-2 antigen variants of all samples of convalescent unvaccinated subjects. The legend of the fingerprint diagram is shown in the box at the bottom of the figure. The left side reports the reference quantification of IgA fraction in terms of GU$_{IgA}$ of three WT antigens, as indicated. The radar chart reports the values of GU$_{IgA}$ for the WT, alfa, gamma, delta and omicron RBD variants. The thick black contour line and the dashed contour line indicate GU$_{IgA}$ = $10^{-2}$ $s^{-1}$ and GU$_{IgA}$ = $0.5 \cdot 10^{-2}$ $s^{-1}$ , respectively.

#### 1.9 Supplementary Figure S7

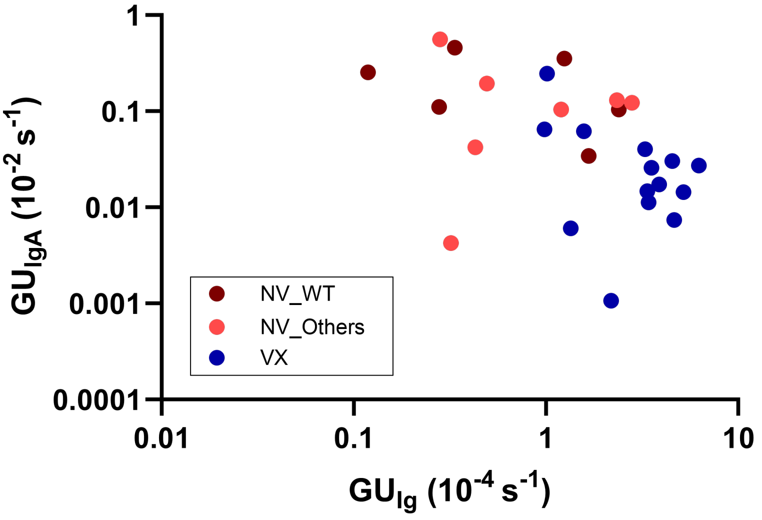


**Figure S7.** Scatter plot for IgA fraction versus Ig levels for all RBD variants. The average values of $GU_{IgA}$ measured for all RBD variants are plotted as a function of the average values of GUIg measured for all RBD variants. The colour of the data point refers to vaccinated subjects (blue) and unvaccinated convalescent subjects infected with WT variant (dark red) or one of the other variants (light red).

#### 1.10 Supplementary Figure S8

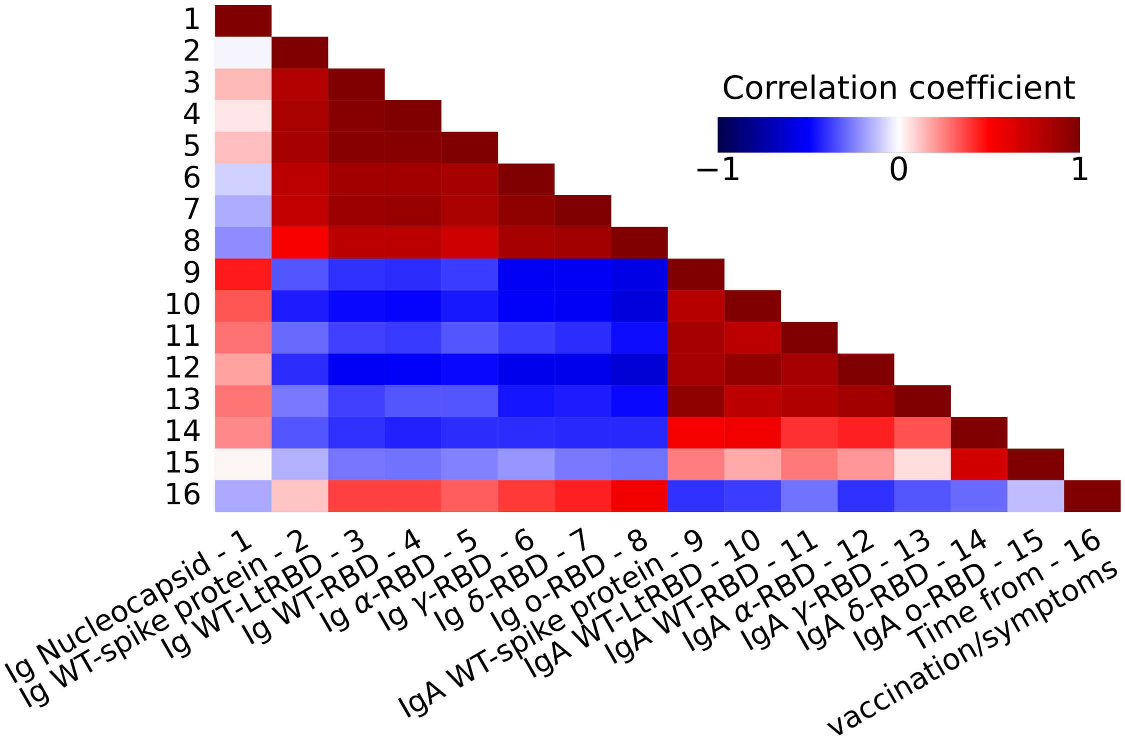


**Figure S8.** Correlation matrix between all Ig and IgA quantification parameters, expressed by GU$_{Ig}$ and GU$_{IgA}$, respectively. The correlation matrix also includes the time (days) passed after vaccination or infection. The Figure shows that Ig and IgA quantification are anti-correlated for all variants, meaning that high levels of total Ig is often associated to low levels if IgA and vice versa. Additionally, the levels of total Ig are all correlated among them, whereas the correlations among the IgA levels are more heterogeneous, being GU$_{IgA}$ for delta and omicron variants weakly correlated with the WT, alfa and gamma variants. Similarly, anti-nucleocapsid Ig levels are weakly or anti-correlated with anti-RBD and anti-spike protein Ig, but the correlation with IgA levels is larger for all variants except for omicron. The time from vaccination or symptoms shows a positive correlation with Ig levels and negative with IgA. This behaviour is ascribed to the rather short interval between infection or vaccination and sample collection of the analysed samples (Supplementary Tables S1 and S2).

#### 1.11 Supplementary Figure S9

a

```
WT-RBD          RVQPTESIVRFPNITNLCPFGEVFNATRFASVYAWNRKRISNCVADYSVLYNSASFSTFK     60
alfa-RBD        RVQPTESIVRFPNITNLCPFGEVFNATRFASVYAWNRKRISNCVADYSVLYNSASFSTFK     60
gamma-RBD       RVQPTESIVRFPNITNLCPFGEVFNATRFASVYAWNRKRISNCVADYSVLYNSASFSTFK     60
delta-RBD       RVQPTESIVRFPNITNLCPFGEVFNATRFASVYAWNRTRISNCVADYSVLYNSASFSTFK     60
omicron-RBD     RVQPTESIVRFPNITNLCPFDEVFNATRFASVYAWNRKRISNCVADYSVLYNLAPFFTFK     60
                *******************.****************.*************** * * ***

WT-RBD          CYGVSPTKLNDLCFTNVYADSFVIRGDEVRQIAPGQTGKIADYNYKLPDDFTGCVIAWNS     120
alfa-RBD        CYGVSPTKLNDLCFTNVYADSFVIRGDEVRQIAPGQTGKIADYNYKLPDDFTGCVIAWNS     120
gamma-RBD       CYGVSPTKLNDLCFTNVYADSFVIRGDEVRQIAPGQTGKIADYNYKLPDDFTGCVIAWNS     120
delta-RBD       CYGVSPTKLNDLCFTNVYADSFVIRGDEVRQIAPGQTGKIADYNYKLPDDFTGCVIAWNS     120
omicron-RBD     CYGVSPTKLNDLCFTNVYADSFVIRGDEVRQIAPGQTGNIADYNYKLPDDFTGCVIAWNS     120
                **************************************:*********************

WT-RBD          NNLDSKVGGNYNYLYRLFRKSNLKPFERDISTEIYQAGSTPCNGVEGFNCYFPLQSYGFQ     180
alfa-RBD        NNLDSKVGGNYNYLYRLFRKSNLKPFERDISTEIYQAGSTPCNGVEGFNCYFPLQSYGFQ     180
gamma-RBD       NNLDSKVGGNYNYRYRLFRKSNLKPFERDISTEIYQAGSKPCNGVEGFNCYFPLQSYGFQ     180
delta-RBD       NNLDSKVGGNYNYLYRLFRKSNLKPFERDISTEIYQAGSTPCNGVKGFNCYFPLQSYGFQ     180
omicron-RBD     NKLDSKVSGNYNYLYRLFRKSNLKPFERDISTEIYQAGNKPCNGVAGFNCYFPLRSYSFR     180
                *:*****.***** ************************..***** ********:**.*:

WT-RBD          PTNGVGYQPYRVVVLSFELLHAPATVCGPKKSTNLVKNKCVNF     223
alfa-RBD        PTYGVGYQPYRVVVLSFELLHAPATVCGPKKSTNLVKNKCVNF     223
gamma-RBD       PTNGVGYQPYRVVVLSFELLHAPATVCGPKKSTNLVKNKCVNF     223
delta-RBD       PTYGVGYQPYRVVVLSFELLHAPATVCGPKKSTNLVKNKCVNF     223
omicron-RBD     PTYGVGHQPYRVVVLSFELLHAPATVCGPKKSTNLVKNKCVNF     223
                ** ***:************************************
```

b

| | WT-RBD | alfa-RBD | gamma-RBD | delta-RBD | omicron-RBD |
|---|---|---|---|---|---|
| **WT-RBD** | 0 | 1 | 3 | 2 | 15 |
| **alfa-RBD** | 1 | 0 | 2 | 3 | 14 |
| **gamma-RBD** | 3 | 2 | 0 | 5 | 16 |
| **delta-RBD** | 2 | 3 | 5 | 0 | 16 |
| **omicron-RBD** | 15 | 14 | 16 | 16 | 0 |

**Figure S9.** Amino acid sequence alignment of SARS-CoV2 RBD variants. (a) Alignment of 223 consecutive amino acids residues from S protein affected by mutations: WT-RBD (Wuhan-Hu-1) Sino Biological Beijing,P.R.China (cod. 40592-V08H); alfa-RBD (B.1.1.7) (N501Y) Sino Biological (cod. 40592-V08H82); gamma-RBD (P.1/ P.1.1/ P.1.2) (K417T, E484K, N501Y) Sino Biological (cod. 40592-V08H86); delta-RBD (B.1.617.2) (L452R, T478K) Sino Biological (cod. 40592-V08H90); omicron-RBD (B.1.1.529) (G339D, S371L, S373P, S375F, K417N, N440K, G446S, S477N, T478K, E484A, Q493R, G496S, Q498R, N501Y, Y505H). Asterisk, colon and period symbols indicate the positions which have a single, fully conserved residue (*), or a conservation between groups of strongly similar properties (:), or a conservation between groups of weakly similar properties (.), respectively. (b) Number of amino acids by which the RBD protein sequences differ.

#### 1.12 Supplementary Figure S10

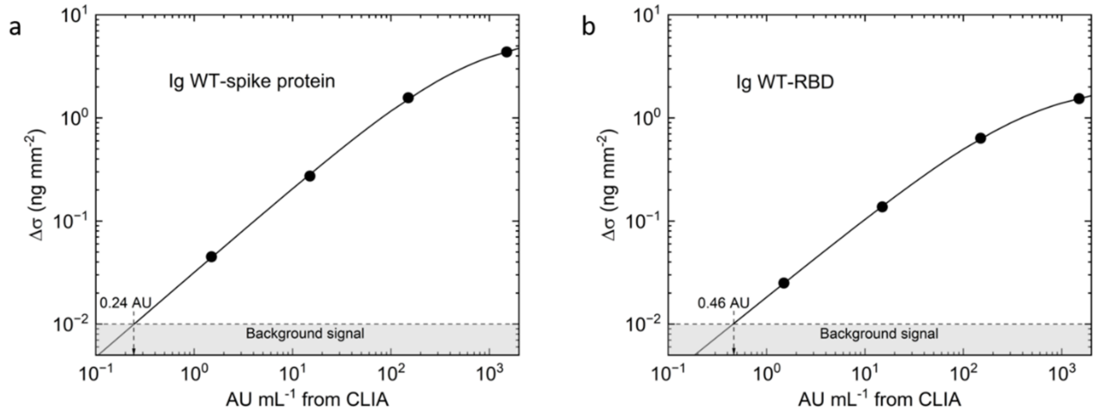


**Figure S10.** Calibration and limit of detection of the RPI antigen micro-array. Surface density of immunoglobulins binding on spots of (a) WT-spike protein and (b) WT-RBD measured by RPI for sample VX14 as a function of the amount of IgG antibodies quantified by Chemiluminescence Enzyme Immunoassays (CLIA kit DiaSorin LIAISON SARS-CoV-2 S1/S2 IgG) expressed as Arbitrary Units per milliliter (AU $mL^{-1}$). The black curves represent fits to the data point with the function $\Delta\sigma = \Delta\sigma_\infty/[1+(K_D/c)^p]$, where $\Delta\sigma_\infty$ is the asymptotic surface density that saturates the surface binding sites, $K_D$ is the equilibrium dissociation constant, c is the IgG concentration measured by CLIA. The values of the parameters obtained from the fit are: $\Delta\sigma_\infty$= 6.6 ng $mm^{-2}$ (a) and $\Delta\sigma_\infty$ = 2.2 ng $mm^{-2}$ (b); $K_D$ = 650 AU $mL^{-1}$ (a) and $K_D$ = 495 AU $mL^{-1}$ (b); p = 0.82 (a) and p = 0.77 (b). The horizontal dashed lines represent the background signal obtained as the maximum non-specific binding response on SARS-CoV-2 antigen spots when plasma sample acquired before the Covid-19 pandemic is added in the measuring cartridge. The limit of detection is indicated by the arrows.

#### 1.13 Supplementary Figure S11

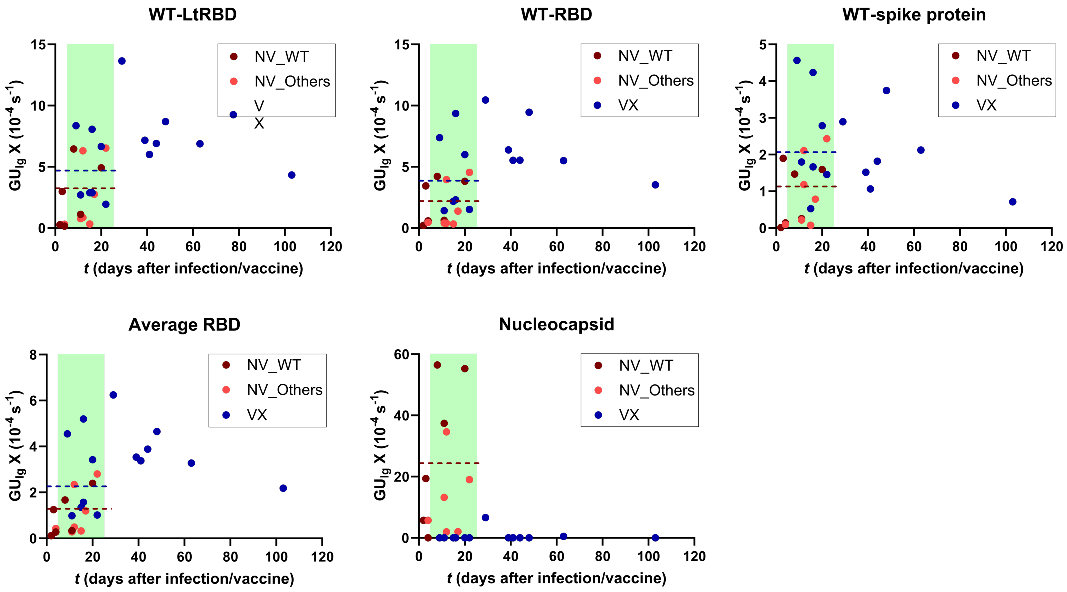


**Figure S11.** Dependence of Ig level on the time after $2^{nd}$ vaccine dose or from the onset of symptoms. The reported values of $GU_{Ig}$ correspond to antibodies binding on spots of WT-LtRBD (a), WT-RBD (b), WT-spike protein (c), all spots of RBD variants (d) and spots of nucleocapsid (e). The colour of the data point refers to Ig levels of vaccinated subjects as a function of days after the $2^{nd}$ dose of vaccine (blue dots) or of unvaccinated subjects infected with WT variant (dark red) or other variants (light red) as a function of days from the onset of symptoms. The green band corresponds to the time window in which vaccinated and unvaccinated samples overlap. The dotted lines are the average values for vaccinated (blue) and all unvaccinated (red) subjects in the overlapping region. It is well known that serum immunoglobulin levels targeting SARS-CoV-2 progressively increase after a few days from vaccination or infection and then tend to decrease in weeks or months Peterhoff et al. (2020) Gaebler et al. (2021). The results are consistent with this behaviour, although the limited time range of the analysed samples does not allow estimating a significant time dependence of the measured parameters. Notably, despite the fact that the samples collected from vaccinated subjects have a larger range of time after the exposure to the virus antigens relative to convalescent subjects, the observations of larger Ig levels and smaller IgA levels for the vaccinated subjects are confirmed also comparing samples collected within similar time ranges.

#### 1.14 Supplementary Figure S12

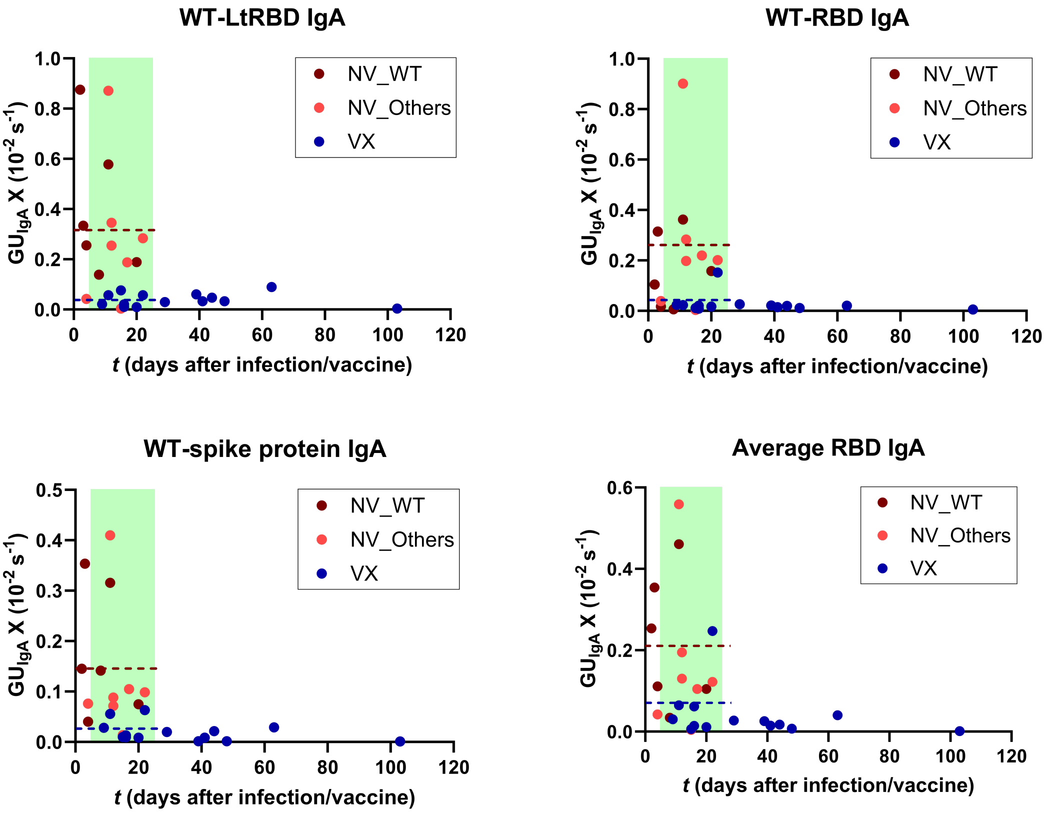


**Figure S12.** Dependence of IgA level on the time after $2^{nd}$ vaccine dose or from the onset of symptoms. The reported values of $\mathrm{GU}_{IgA}$ correspond to antibodies binding on spots of WT-LtRBD (a), WT-RBD (b), WT-spike protein (c), and all spots of RBD variants (d). The colour of the data point refers to IgA levels of vaccinated subjects as a function of days after the $2^{nd}$ dose of vaccine (blue dots) or of unvaccinated subjects infected with WT variant (dark red) or other variants (light red) as a function of days from the onset of symptoms. The green band corresponds to the time window in which vaccinated and unvaccinated samples overlap. The dotted lines are the average values for vaccinated (blue) and all unvaccinated (red) subjects in the overlapping region.

### 2 SUPPLEMENTARY NOTE

#### 2.1 Supplementary Note S1

Principal component analysis (PCA) was performed on the whole data set of Ig and IgA data. PCA enabled determining linear orthogonal combinations of the parameters (in this case the Ig and IgA quantification), named PC1, PC2, PC3 etc. that maximally spread out the value among the 27 subjects, hence carrying the largest amount of information on the data. Accordingly, the variance between the value of PC1 computed on the set of subjects is the largest among any other combination of $GU_{Ig}$ and $GU_{IgA}$ values. PC1, PC2 and PC3 are obtained by multiplying the 15 $GU_{Ig}$ and $GU_{IgA}$ values by optimized arrays of coefficients that are reported in Figure 4c. PCA was made with a Python (v. 3.9.2) routine, using the machine learning library scikit-learn (v. 1.2.0). sklearn.decomposition.PCA() class performed a linear dimensionality reduction through Singular Value Decomposition (SVD). Multidimensional were rescaled to unit variance and centred around zero prior to processing using the .fit_transform(method) of the sklearn.preprocessing.StandarScaler() class.

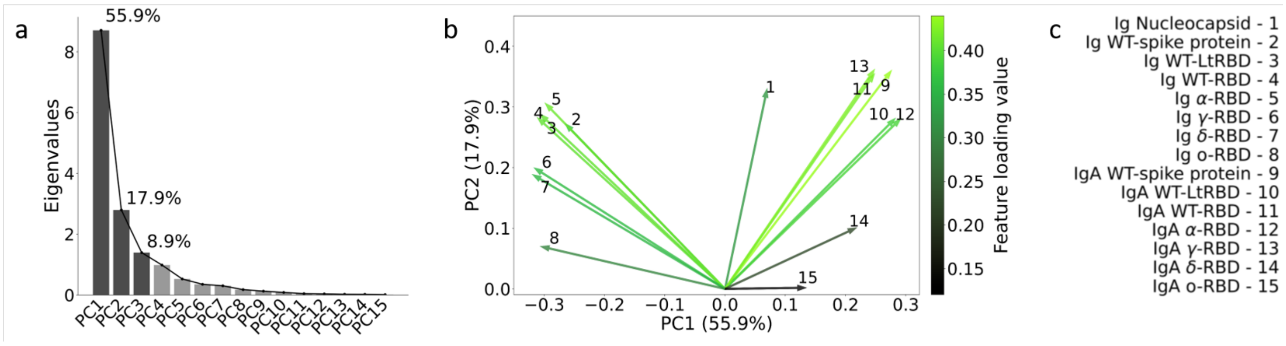


**Figure S13.** Principal component analysis of antibody fingerprints. (a) Scree plot of PCA, which visually presents the eigenvalues and their corresponding explained variance for the first three components. The eigenvalues represent the amount of variance captured by each component, with higher eigenvalues indicating components that explain more variability in the data. The explained variance (reported as percentage), shown alongside the eigenvalues, quantifies the proportion of total variance accounted for by each respective component. The scree plot aids in determining the significance and contribution of each component in summarizing the dataset's variability and assessing the optimal number of components to retain in the analysis. (b) The arrows centered in the origin represent the projection of each feature on the PC1-PC2 plane. The color of the arrows represent the loading value of each feature, according to the color bar. (c) Antibody type and antigen spot associated to each feature of panel b.
